# Comparative Analysis of Machine Learning Models vs. Traditional Clinical Calculators for Cardiovascular Risk Prediction

**DOI:** 10.64898/2026.06.11.26355407

**Authors:** Nicolás Arango Plaza, Gladys Elena Salcedo Echeverry, Ailan Farid Arenas-Soto

**Affiliations:** Doctorado en Ciencias Biomédicas, Universidad del Quindío. Armenia-Quindío, Colombia; Facultad de Educación, Programa de Licenciatura en Matemáticas, Universidad del Quindío. Armenia-Quindío, Colombia; Facultad de Ciencias de la Salud, Universidad del Quindío. Armenia-Quindío, Colombia; Grupo de Asesoría e Investigación en Estadística. Universidad del Quindío. Armenia-Quindío, Colombia

## Abstract

**Background:** Cardiovascular diseases (CVD) remain the leading global cause of mortality, responsible for approximately 31% of all deaths worldwide in 2021. Traditional risk calculators, including Framingham, ASCVD, SCORE, and SCORE2, have long constituted the cornerstone of primary prevention strategies; however, they were derived predominantly from high-income European and North American populations, thereby limiting their predictive accuracy in diverse epidemiological contexts, particularly among Hispanic/Latino communities. Machine learning (ML) offers an alternative to capture the non-linear interactions inherent in biomedical data.

**Objective:** The present study develops and validates ML-based models for cardiovascular mortality prediction using the National Health and Nutrition Examination Survey (NHANES) 1999–2018 dataset, and systematically compares their discriminative performance against eleven conventional clinical CVD risk calculators.

**Materials and Methods:** A dedicated software platform, “CardioPrediQ,” was designed to integrate multiple CVD calculators with ML-based risk assessment. A cohort of 12,847 participants with 16 predictor variables was derived from NHANES. Six algorithms (Logistic Regression, Cox Proportional Hazards, Gradient Boosting, AdaBoost, Random Forest, and Extra Trees) were trained in combination with six class-balancing strategies, yielding 36 model configurations. All models were trained on a stratified 70/30 split and calibrated using the Saerens prior probability adjustment method. Performance was evaluated using AUC-ROC, sensitivity, specificity, F1-score, and a weighted composite score. DeLong’s test was employed to assess the statistical significance of AUC differences between the best-performing ML model and each conventional calculator.

**Results:** Gradient Boosting with 2:1 oversampling and Saerens calibration achieved the best overall performance (AUC = 0.8934; composite score = 0.7904), outperforming all traditional calculators in composite ranking. The top six positions were occupied exclusively by ML and statistical models. The mean age of cardiovascular decedents was 67.43 years compared with 47.74 years among survivors. DeLong’s test confirmed statistical superiority over six traditional CVD calculators (p < 0.05), whereas the difference against the top-performing calculators (ASCVD, HEARTS Caribbean, ASCVD Colombia, SCORE2, HEARTS North America) did not reach statistical significance. Age dominated feature importance at 41.2% relative weight, followed by systolic blood pressure (18.7%). Saerens calibration reduced the Brier score from 0.1286 to 0.1158, substantially improving probability calibration.

**Conclusions:** ML models demonstrated superior composite performance over traditional calculators. The statistical equivalence with the highest-performing conventional calculators in the NHANES cohort is context-dependent and validates the methodological pipeline. The CardioPrediQ platform addresses the critical need for integrated, scalable CVD risk assessment tools, which is particularly relevant for Latin American populations where calculator validation remains limited. These findings support the integration of calibrated ML-based risk prediction into clinical practice while underscoring the importance of probability calibration for informed clinical decision-making.

## 1. Introduction

Cardiovascular diseases (CVD) constitute the most formidable challenge to global public health in the twenty-first century. In 2021, approximately 60 million deaths occurred worldwide, of which 31% were attributable to cardiovascular causes, thereby sustaining CVD as the leading cause of mortality across all regions [1]. This burden is particularly acute in low and middle-income countries (LMICs), where CVD mortality rates often exceed those of high-income nations owing to limited access to preventive care, delayed diagnosis, and inadequate risk stratification tools [2]. In Latin America and the Caribbean alone, CVD mortality reached 188 per 100,000 males and 132 per 100,000 females in 2021, with ischaemic heart disease persisting as the single largest cause of death [2]. The cornerstone of primary CVD prevention is the accurate estimation of individual cardiovascular risk, which informs clinical decisions regarding pharmacological intervention, lifestyle modification, and follow-up intensity. Over the past four decades, a family of risk prediction tools has been developed for this purpose, including the Framingham Risk Score [3], the Systematic COronary Risk Evaluation (SCORE) [4], its updated version SCORE2 [5], the Pooled Cohort Equations for Atherosclerotic Cardiovascular Disease (ASCVD) [6], the Prospective Cardiovascular Münster Study (PROCAM) [7], the REGICOR score [8], and the HEARTS in the Americas initiative of the World Health Organization and Pan American Health Organization (WHO/PAHO) [9]. However, a fundamental barrier to the widespread applicability of these traditional risk calculators is that they were derived and validated predominantly in populations from high-income European or North American countries, and their calibration deteriorates substantially when applied to populations with different risk factor distributions, genetic backgrounds, or environmental exposures [10, 11]. For instance, the PURE-Colombia study demonstrated that all six evaluated risk models exhibited adequate discrimination but poor calibration among Colombian adults, even with recalibration [11]. Similarly, a large-scale validation in 112,262 Mexican adults confirmed that all models required region-specific recalibration to achieve acceptable risk estimation [12]. The strategic convergence of high-fidelity digitized clinical data and machine learning (ML) offers a promising alternative to traditional linear survival regression models. Unlike conventional tools, ML algorithms can navigate the high-dimensional, non-linear interactions characteristic of biomedical data. Weng et al. (2017) demonstrated that ML significantly improved CVD risk prediction over QRISK2 and Framingham using routine clinical data from 378,256 patients [13]. More recently, Ahiduzzaman and Hasan (2025) showed that XGBoost achieved the highest accuracy (0.8216) for CVD prediction using NHANES 2017–2023 data [14]. In Latin American contexts, Reátegui et al. (2025) applied ensemble ML with oversampling methods to cardiovascular risk estimation in an Ecuadorian population [15]. The American Heart Association has recognized the potential of advanced statistical methods with the release of the PREVENT equations, which incorporate kidney and metabolic measures beyond traditional risk factors [16]. A critical challenge in applying ML to clinical CVD prediction is the severe class imbalance inherent in cardiovascular cohort data, wherein survivors vastly outnumber events. Various resampling strategies, including SMOTE [17], random oversampling, undersampling, and Tomek Links, have been proposed to mitigate this imbalance; however, their impact on model calibration in clinical prediction remains controversial [18]. The National Health and Nutrition Examination Survey (NHANES), administered by the Centers for Disease Control and Prevention (CDC), provides a nationally representative sample of the United States population with longitudinal mortality follow-up through linkage with the National Death Index (NDI). NHANES is particularly valuable for the present research for three reasons: first, it serves as the primary data source for several of the compared calculators (notably ASCVD and Framingham), thereby enabling direct comparison; second, its substantial Hispanic population subgroup allows investigation of South American cardiovascular prediction assessments; and third, its comprehensive clinical variables enable the simultaneous implementation and evaluation of multiple international calculators. In this study, we designed a software platform termed “CardioPrediQ” that integrates twelve international CVD calculators with ML-based risk prediction, intended to serve as a clinical decision support tool for both research and primary care settings. With this tool, we developed and validated a systematic comparison of 36 ML model combinations against eleven conventional cardiovascular risk calculators using the NHANES 1999–2018 dataset. We hypothesized that machine learning models trained on NHANES data with appropriate class-balancing strategies and probability calibration would achieve superior composite discriminative performance compared to traditional clinical cardiovascular risk calculators.

## 2. Materials and Methods

### Data Source and Study Population

This study utilized data from the National Health and Nutrition Examination Survey (NHANES), administered by the CDC’s National Center for Health Statistics. The analysis incorporated biennial survey cycles from 1999–2000 through 2017–2018, with mortality follow-up through December 2019 via linkage with the National Death Index. The outcome variable was cardiovascular mortality, defined as death from heart disease or cerebrovascular disease. Surviving participants required at least 120 months of documented follow-up.

### Variable Selection

From approximately 5,000 variables available in the combined NHANES database, 14 clinical variables were initially selected to match the inputs required by the cardiovascular risk calculators implemented in CardioPrediQ. These comprised 16 predictor variables: Age, Sex, Weight, Height, Systolic Blood Pressure (SBP), Total Cholesterol, HDL, LDL, Triglycerides, Diabetes, Smoking Status, Antihypertensive Treatment, Race (ASCVD classification), and Family History of Premature Cardiovascular Disease (FHPCVD). Additionally, seven engineered features were created for the expanded input block (BAE), including Body Mass Index (BMI), Non-HDL cholesterol, Atherogenic Index, Total Cholesterol-to-HDL ratio, and age × SBP interaction terms, yielding 21 input variables for tree-based models.

### Data Preprocessing

A nine-stage cleaning pipeline was implemented in R, reducing the raw database from approximately 230,000 records to the final analytical sample of 12,847 participants (S1 Appendix). Key preprocessing decisions included the following: the exclusion of participants not classifiable as Non-Hispanic White or Non-Hispanic Black owing to the ASCVD calculator’s binary race requirement; the recovery of 40,425 missing LDL values using the Friedewald formula (LDL = TC − HDL − TG/5) conditioned on triglycerides < 400 mg/dL, employing original values of TC, HDL, and TG; the reclassification of borderline diabetes (code 3) as positive in accordance with the ACC/AHA 2013 guidelines; and the application of the IQR × 3 criterion for outlier elimination, which is considerably more permissive than the standard IQR × 1.5 threshold to avoid discarding genuinely extreme but valid values. The final analytical sample comprised 12,847 participants with a class imbalance ratio of 5.29:1 (84.1% alive, 15.9% deceased), consistent with the expected prevalence in cardiovascular cohort studies.

### Train/Test Partition and Class Balance

The dataset was partitioned using a stratified 70/30 split (random_state = 42), preserving the exact 15.9% event prevalence in both subsets: training set n = 8,992 (7,563 alive, 1,429 deceased); test set n = 3,855 (3,242 alive, 613 deceased). The test set remained entirely unmodified throughout all training and evaluation processes. Six class-balancing strategies were evaluated exclusively on the training set: original sample (ratio 5.29:1); random oversampling 1:1; random oversampling 2:1; random oversampling 3:1; random undersampling 1:1; and SMOTE with k = 5 nearest neighbors. The Cox Proportional Hazards model required no balancing, as its Breslow partial likelihood natively accommodates differential censoring. Because oversampling artificially inflates the prevalence of events in the training set, raw model outputs may overestimate the true risk. Because oversampling alters the empirical class distribution of the training data, raw posterior probabilities produced by the classifiers may not reflect the true prevalence of events in the target population. Therefore, predicted probabilities were corrected using the Saerens prior probability adjustment method, which recalibrates posterior estimates according to the expected deployment prevalence under the assumption of invariant class-conditional feature distributions [19]. A composite performance index was calculated as a weighted combination of discrimination, classification, agreement, and calibration metrics: 40% area under the ROC curve (AUC), 20% sensitivity, 20% F1-score, 10% Cohen’s Kappa, and 10% inverse Brier score (1 − Brier). The composite score was defined as:

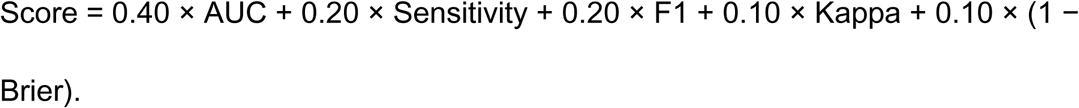

AUC was assigned the highest weight as the most robust metric for comparing models in imbalanced contexts; sensitivity was prioritized because false negatives (i.e., missed high-risk patients) carry greater clinical consequences than false positives; the F1-score penalizes models that sacrifice precision for sensitivity; Cohen’s Kappa measures agreement beyond chance; and the complement of the Brier score assesses probabilistic calibration.

### Comparison with Conventional CVD Risk Calculators

The 36 ML configurations (6 algorithms × 6 balancing strategies) were evaluated and compared against eleven conventional CVD risk calculators: ASCVD Original, ASCVD Colombia, Framingham Original, Framingham Colombia, High SCORE, Low SCORE, SCORE2, HEARTS Caribbean, HEARTS North America, PROCAM, and REGICOR. All calculators were implemented in JavaScript within CardioPrediQ and applied to the test set using each participant’s clinical variables. Discrimination was quantified using AUC-ROC, and DeLong’s test was employed to determine the statistical significance of AUC differences between the best-performing ML model and each conventional CVD risk calculator.

### Software Design: CardioPrediQ

CardioPrediQ is a web application deployed as a self-contained HTML file (8,572 lines of HTML + CSS + JavaScript) that integrates twelve cardiovascular risk calculators with ML-based prediction. The application requires no backend infrastructure; all processing occurs in the user’s browser, ensuring that patient data never leaves the local device, a critical privacy consideration for clinical software. The architecture follows a monolithic single-page application (SPA) pattern without a reactive framework, organized into five logical layers: 1) Presentation (HTML5 + CSS3 with a light/dark theme system using CSS Custom Properties); 2) Visual Design (glassmorphism, fluid typography with clamp(), color-mix() for theme adaptation); 3) UI Logic (panel state management, form validation, real-time Friedewald LDL calculation); 4) Calculation Engine (twelve calculator algorithms encoded in Base64, decoded via atob() and injected via eval()); and 5) Export (clinical PDF generation via jsPDF, Excel export via SheetJS, multi-patient batch mode). The calculator engine includes three models validated in Colombia: 1) Framingham Colombia, which applies a 0.75 multiplier derived from the PURE-Colombia study; 2) ASCVD Colombia, recalibrated with factors of 0.54 (women) and 0.28 (men) from the PURE-Colombia cohort [11]; and 3) HEARTS Colombia (region hoa10, Central Latin America), employing WHO/PAHO risk tables for the most appropriate geographic region [9]. Additionally, nine international calculators were integrated (Framingham, ASCVD, SCORE, SCORE2, SCORE2-OP, PROCAM, REGICOR, and HEARTS with 21 selectable global regions). A distinctive feature is the interactive SVG anatomical figure that provides visual risk feedback: upon calculation, the figure’s color changes dynamically according to the risk category (teal for low, amber for moderate, orange for high, red for very high, burgundy for critical), accompanied by a pulse animation confirming the calculation. The Friedewald equation is automatically applied when LDL is missing (LDL = TC − HDL − TG/5), with a precision warning when TG ≥ 400 mg/dL. Each calculator’s primary bibliographic reference is displayed in APA format upon selection. The software was registered with Colombia’s National Directorate of Copyright (DNDA), Contract No. 246/2024, and is publicly available at https://nicolasarangoplaza.github.io/CardioPrediQ/

## 3. Results

From approximately 5,000 variables available in the combined NHANES database, 14 clinical variables were initially selected to match the inputs required by the cardiovascular risk calculators, which, together with two additional derived variables, yielded 16 predictor variables. The nine-stage cleaning pipeline reduced the raw database from approximately 230,000 records to the final analytical sample of 12,847 participants (S1 Appendix).

### Exploratory Analysis

The analytical cohort comprised 12,847 participants (mean age 50.87 ± 14.48 years, range 30 to 80). The cohort was 47.1% male (n = 6,047) and 52.9% female (n = 6,800). Under the ASCVD binary race classification, the sample included Non-Hispanic White participants (including Hispanic/Latino and North American White individuals; n = 8,831, 68.7%) and Non-Hispanic Black participants (n = 4,016, 31.3%); a separate ethnicity classification identified 31.2% of participants as Hispanic/Latino. Baseline cardiovascular risk factors included: hypertension treatment (24.2%, n = 3,113), current smoking (51.0%, n = 6,555), diabetes (9.8%, n = 1,255), and family history of premature cardiovascular disease (FHPCVD) (15.7%, n = 2,013). Mean total cholesterol was 204.41 ± 40.55 mg/dL, HDL 53.63 ± 15.94 mg/dL, LDL 123.52 ± 36.56 mg/dL, triglycerides 131.75 ± 73.13 mg/dL, and systolic blood pressure (SBP) 125.13 ± 17.85 mmHg. Mean weight was 82.65 ± 19.39 kg and mean height 170.76 ± 9.42 cm (S1 Appendix). During follow-up (median 14.2 years, IQR 9.1–17.8), 2,042 cardiovascular deaths occurred (15.9% of the cohort). The mean age of cardiovascular decedents was 67.43 years compared with 47.74 years among survivors (P25 = 38 years, P75 = 62 years). The 10-year cardiovascular mortality rate was 8.4%, with significant variation by age stratum: 3.1% for 30–44 years, 8.7% for 45–64 years, and 18.2% for 65–74 years (S1 Appendix).

Principal Component Analysis (PCA) on the 16 predictor variables demonstrated that eight components were required to explain 81.3% of the total variance, confirming that cardiovascular risk is inherently multi-dimensional and that no single variable acts in isolation. The first principal component (PC1), accounting for 16.48% of the variance, represented an anthropometric axis, with sex, weight, and height positively loaded, whereas HDL was negatively loaded. The second principal component (PC2), accounting for 15.21% of the variance, captured a lipidic-hypertensive axis with Total Cholesterol, LDL, and SBP. Critically, no linear separation between alive and deceased participants was observed in the PC1–PC2 biplot, confirming the necessity for non-linear modeling approaches (Fig 1).

**Fig 1.**
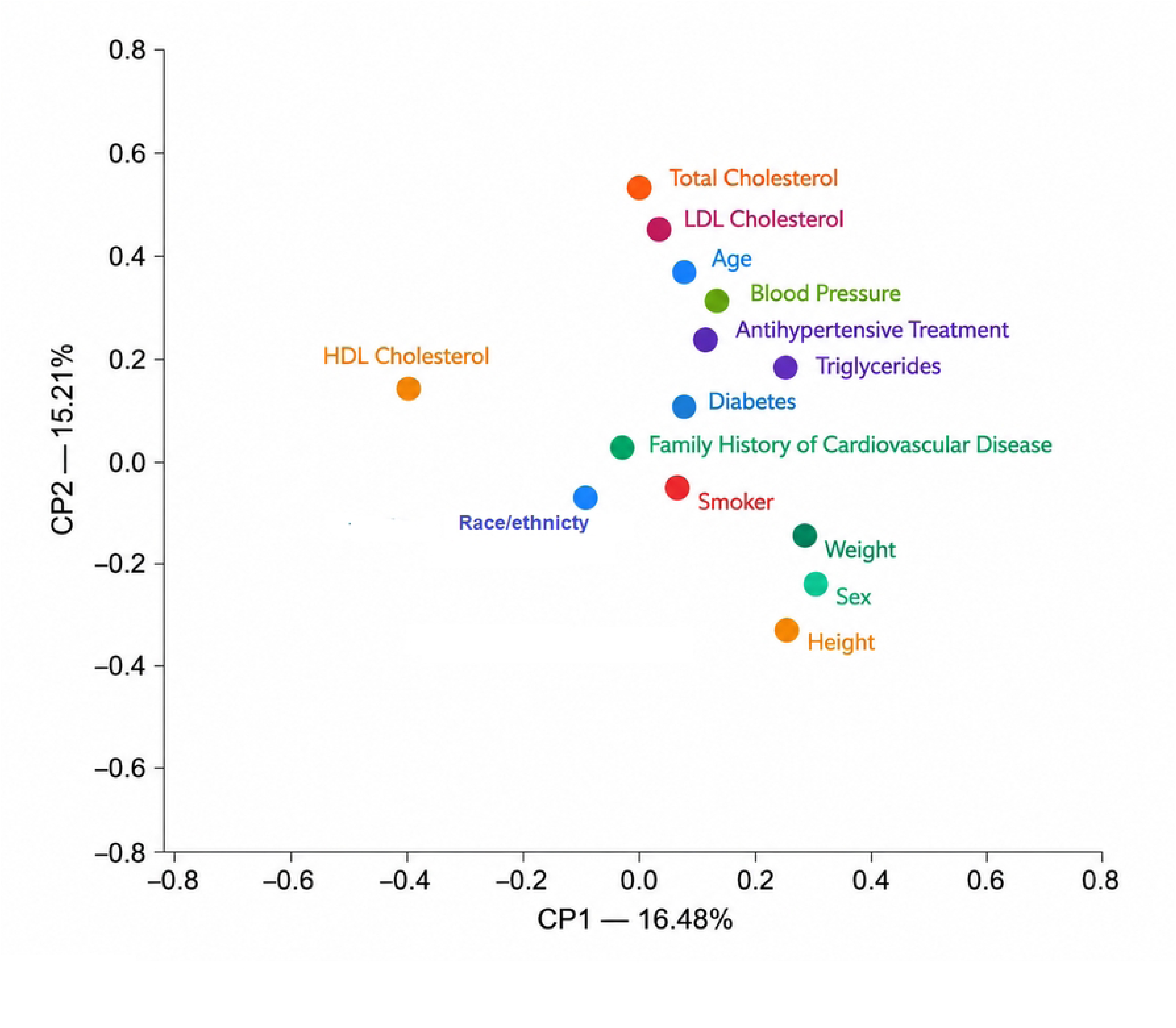
Principal component analysis (PCA) biplot of the CVD predictor variables. PC1 (16.48% variance explained) represents an anthropometric axis; PC2 (15.21% variance explained) represents a lipidic-hypertensive axis. No linear separation between alive and deceased participants is evident.

Age presented the highest individual correlation with vital status (r = 0.50), followed by systolic blood pressure (r = 0.28) and antihypertensive treatment (r = 0.25). The correlation matrix revealed multicollinearity between Total Cholesterol and LDL (r = 0.91) and a moderate negative correlation between HDL and Triglycerides (r = −0.42) (Fig 2). Distribution analysis confirmed that cardiovascular deaths are concentrated at ages above 63 years, with a marked shift toward older age groups among decedents (S1 Appendix).

**Fig 2.**
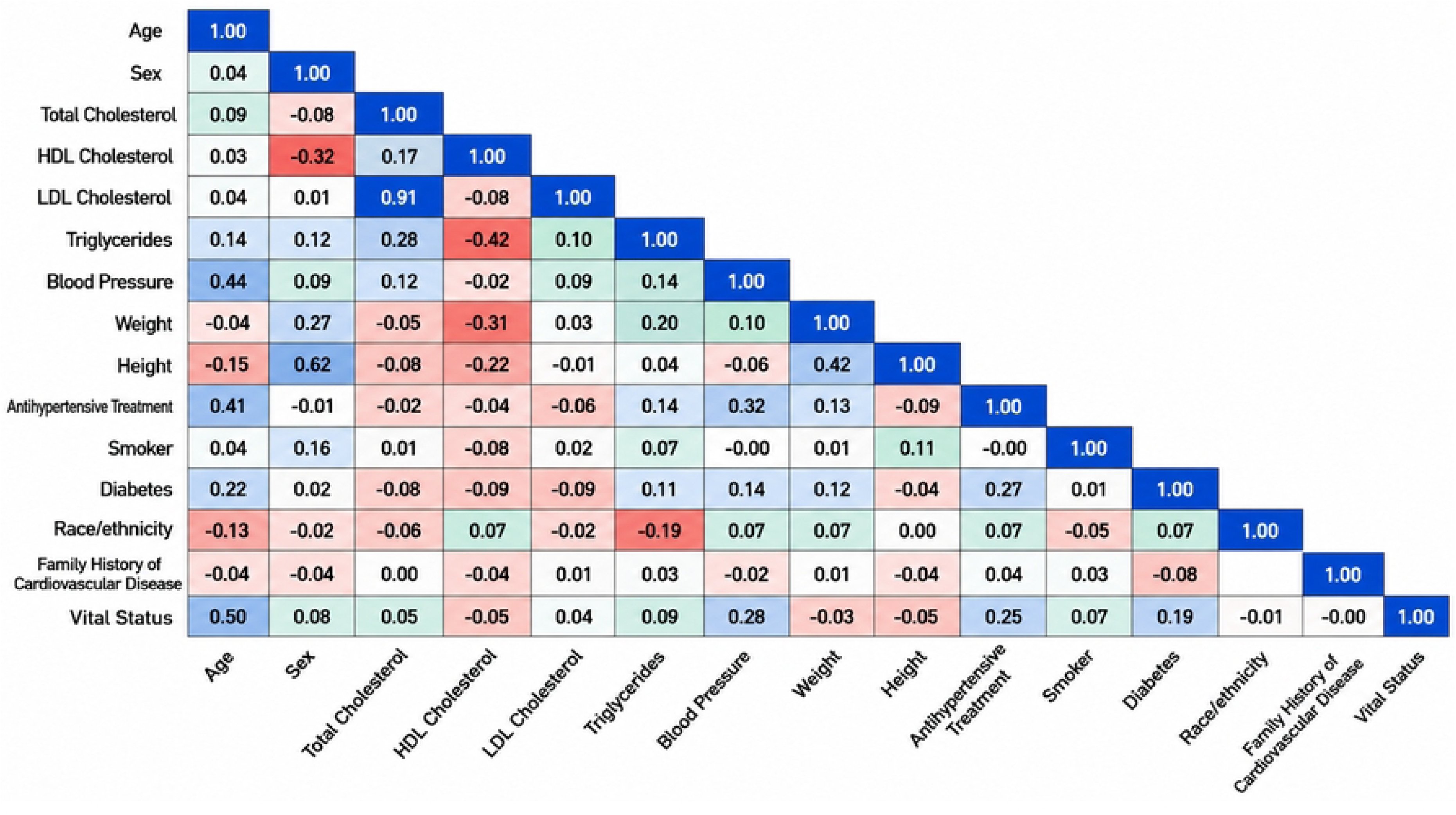
Heatmap correlation matrix of the predictor variables. The strongest positive correlation was observed between Total Cholesterol and LDL (r = 0.91); the strongest negative correlation was between HDL and Triglycerides (r = −0.42). Age showed the highest correlation with vital status (r = 0.50).

### Model Performance

Thirty-six ML configurations (6 algorithms × 6 balancing strategies) were evaluated. The Gradient Boosting Machine (GBM) with Saerens probability calibration and 2:1 oversampling achieved the highest AUC of 0.8934 and the highest composite score of 0.7904. The top six positions in the composite ranking were occupied exclusively by ML and statistical models: GBM (AUC 0.8934, score 0.7904), Logistic Regression with SMOTE (AUC 0.8864, score 0.7840), Extra Trees with 3:1 oversampling (AUC 0.8817, score 0.7801), AdaBoost with 3:1 oversampling (AUC 0.8856, score 0.7785), Random Forest with undersampling (AUC 0.8858, score 0.7750), and Cox Regression (AUC 0.8826, score 0.7746) (Fig 3). The Neural Network with undersampling achieved an AUC of 0.8137, which was the lowest among all models evaluated, suggesting that the architecture (64,32 hidden layers with 1,000 iterations) was suboptimal for the complexity of the prediction task. In contrast, ensemble methods (GBM, AdaBoost, Random Forest, Extra Trees) consistently ranked in the top positions regardless of balancing strategy (Fig 3).

**Fig 3.**
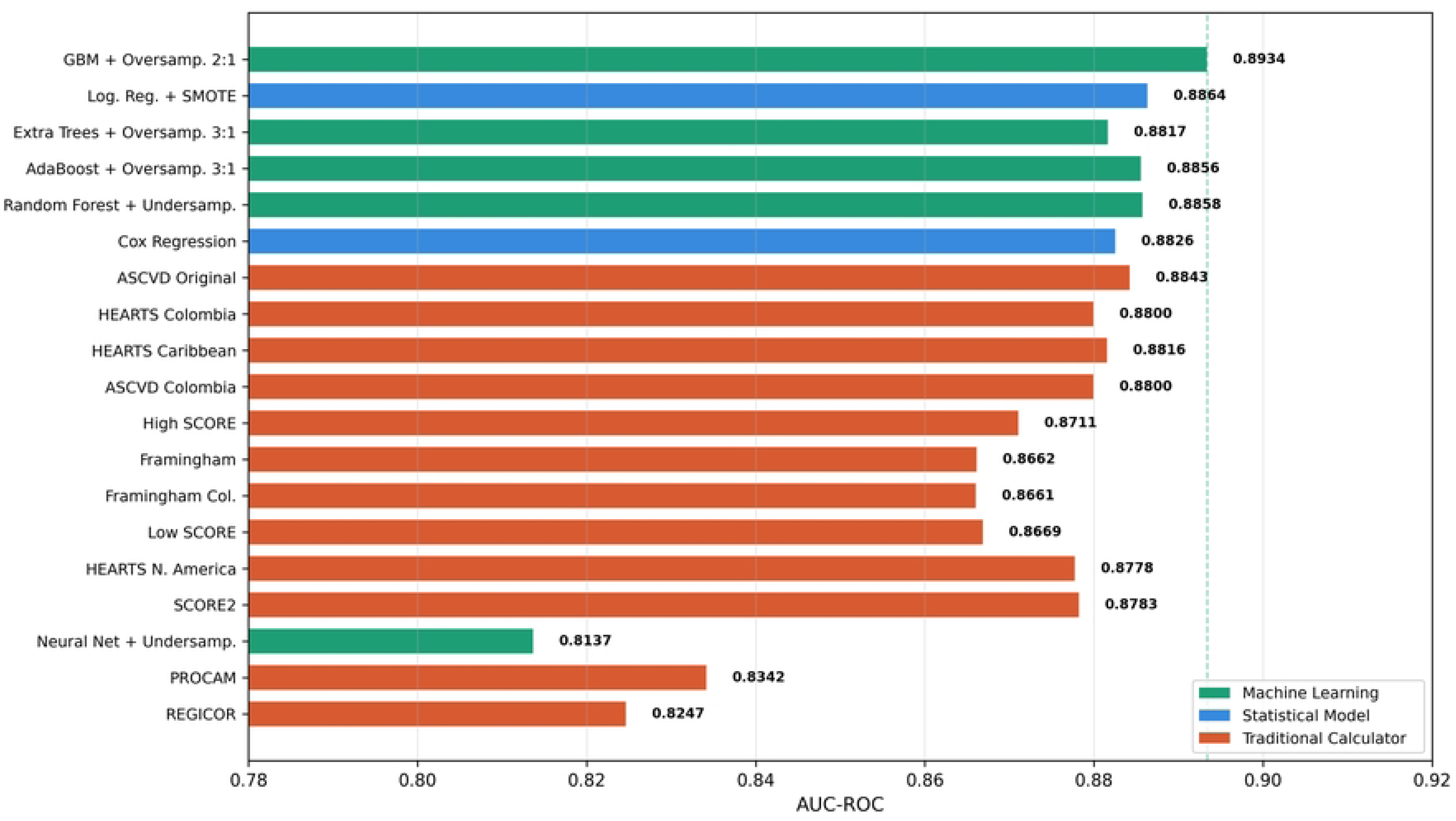
Comparative AUC-ROC across ML models and CVD risk calculators. ML models (green) and statistical models (blue) occupy the top six positions, outperforming traditional CVD risk calculators (orange). The outcome variable was vital status (alive or deceased due to cardiovascular disease).

The effect of class balancing varied across algorithms. Simple oversampling to 2:1 emerged as the optimal strategy for tree-based models, whereas SMOTE with k = 5 provided marginal improvements for Logistic Regression (+0.006 in composite score). Random undersampling, despite discarding 68% of the majority class data, exhibited the lowest variance across model runs, indicating high consistency. No single balancing strategy universally outperformed the others; rather, the interaction between algorithm characteristics and class distribution constituted the primary determinant of performance. The eleven traditional cardiovascular risk calculators exhibited a performance range of AUC-ROC from 0.8843 to 0.8247. The ASCVD Original, derived from US cohorts, achieved the highest AUC among traditional calculators (0.8843), followed closely by HEARTS Caribbean (0.8816), HEARTS Colombia (0.8800), and ASCVD Colombia (0.8800). The REGICOR and PROCAM algorithms exhibited the lowest performance (AUC 0.8247 and 0.8342, respectively), likely reflecting their Spanish and German derivation populations. The relatively narrow range of AUC values (0.82-0.88) across calculators suggests that, in the NHANES population, the derivation population exerts a less pronounced effect than fundamental risk factor relationships (Fig 3).

A comparative analysis of five performance metrics for the top ML models and CVD risk calculators revealed a critical tradeoff. While calculators such as ASCVD Original and HEARTS Caribbean achieved higher sensitivity (0.89–0.95), this advantage came at a substantial cost in specificity (0.59–0.71) and F1-score (0.41–0.52), indicating a tendency toward risk overprediction. In contrast, ML models maintained a more balanced sensitivity-specificity tradeoff: GBM achieved a sensitivity of 0.8515 and a specificity of 0.8167 simultaneously, with an F1-score of 0.6021. Notably, HEARTS North America exhibited the highest sensitivity (0.9804) but the lowest specificity (0.4303) among all models, whereas REGICOR showed the opposite pattern—high specificity (0.9710) but markedly low sensitivity (0.2594) (Fig 4).

**Fig 4.**
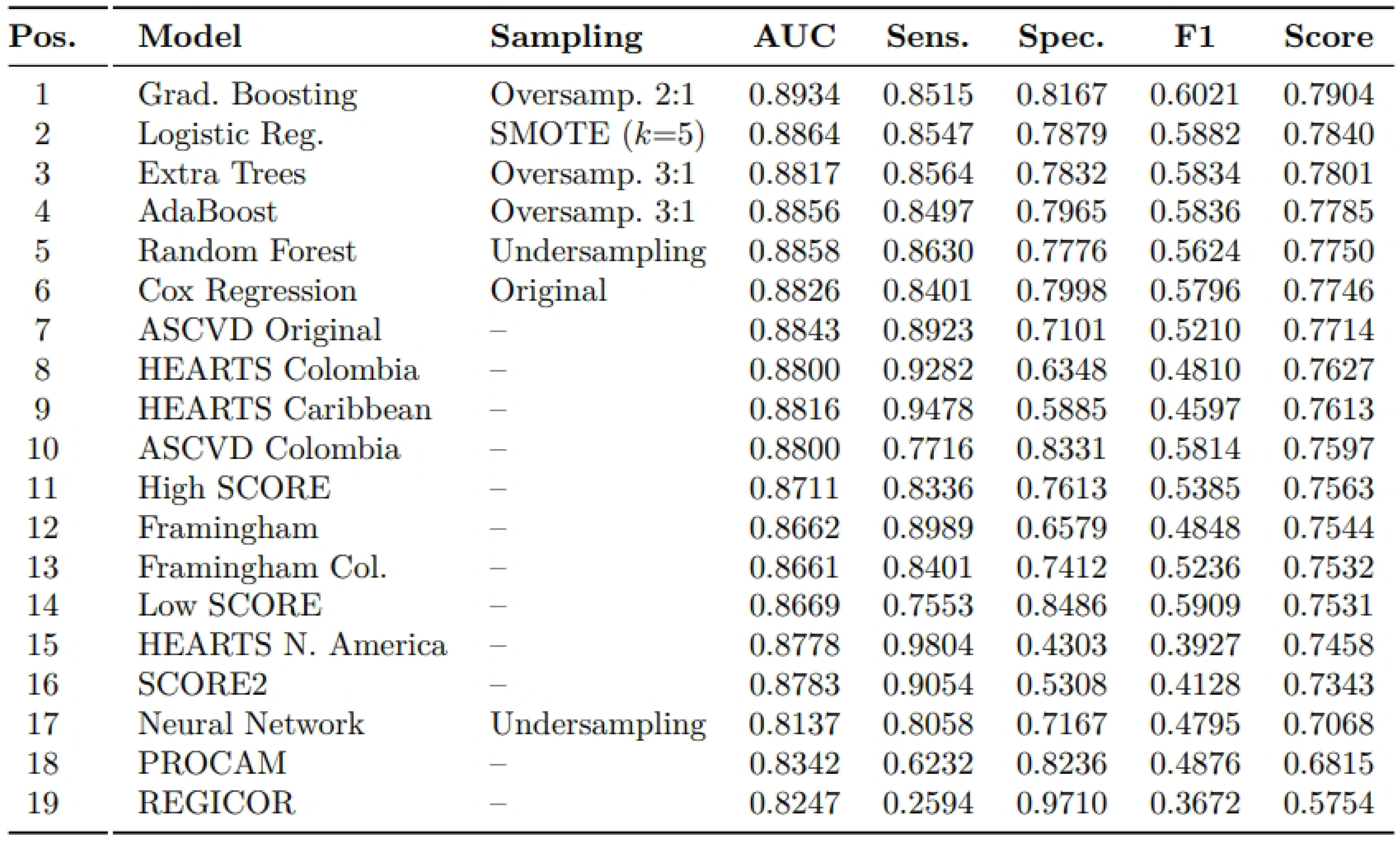
Heatmap correlation matrix illustrating the tradeoff across five performance metrics (AUC-ROC, sensitivity, specificity, F1-score, and composite score). While ASCVD and HEARTS (Caribbean and Colombia) exhibited high sensitivity, their low specificity indicates a tendency toward overprediction of cardiovascular risk. ML models demonstrated more balanced metrics across all five dimensions.

### ML vs CVD Risk Calculators “DeLong Tests”

DeLong’s test was employed to compare the AUC of the best-performing model (GBM + Saerens, AUC 0.8934) against each risk calculator. Statistically significant differences were observed against calculators derived from non-US populations: High SCORE (ΔAUC = 0.0223, p = 0.0238), Low SCORE (ΔAUC = 0.0264, p = 0.0094), Framingham Original (ΔAUC = 0.0272, p = 0.0044), Framingham Colombia (ΔAUC = 0.0272, p = 0.0044), PROCAM (ΔAUC = 0.0591, p < 0.0001), and REGICOR (ΔAUC = 0.0687, p < 0.0001). The difference against ASCVD Original, while numerically observable (+0.0090 AUC), did not reach statistical significance (p = 0.3228). Similarly, the differences against HEARTS Caribbean (ΔAUC = 0.0117, p = 0.2007), ASCVD Colombia (ΔAUC = 0.0134, p = 0.1412), SCORE2 (ΔAUC = 0.0150, p = 0.1121), and HEARTS North America (ΔAUC = 0.0155, p = 0.0966) were not statistically significant, indicating that the top ML model and the highest-performing CVD risk calculators perform comparably in this population (Fig 5).

**Fig 5.**
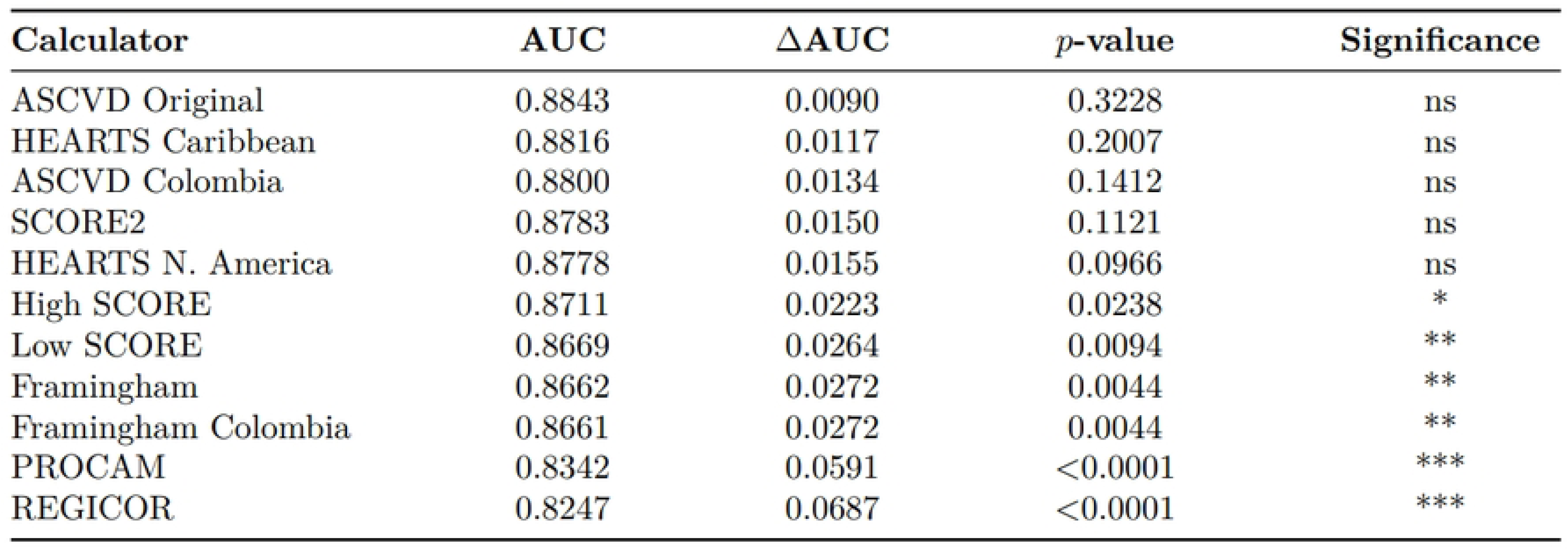
DeLong’s test results comparing GBM + Saerens (AUC = 0.8934) against each traditional CVD risk calculator. Statistical significance: *p < 0.05, **p < 0.01, ***p < 0.001; ns, not significant.

### Feature Importance

Variable importance analysis from the GBM model revealed that age dominated prediction with a relative importance of 0.412 (41.2%), followed by systolic blood pressure (SBP) (0.187, 18.7%), Total Cholesterol (0.094, 9.4%), HDL (0.081, 8.1%), and Smoking (0.063, 6.3%). Together, these five features accounted for 83.7% of the model’s discriminative capacity. Subsequent contributors included Diabetes (0.045, 4.5%), LDL (0.035, 3.5%), BMI (0.028, 2.8%), Sex (0.022, 2.2%), Triglycerides (0.015, 1.5%), Race/ethnicity (0.008, 0.8%), Antihypertensive Treatment (0.005, 0.5%), Family History of CVD (0.003, 0.3%), Weight (0.001, 0.1%), and Height (0.001, 0.1%). Notably, the engineered features (BMI, Non-HDL cholesterol, Atherogenic Index) collectively contributed modestly, suggesting that the primary clinical variables suffice for most of the discriminative capacity (Fig 6).

**Fig 6.**
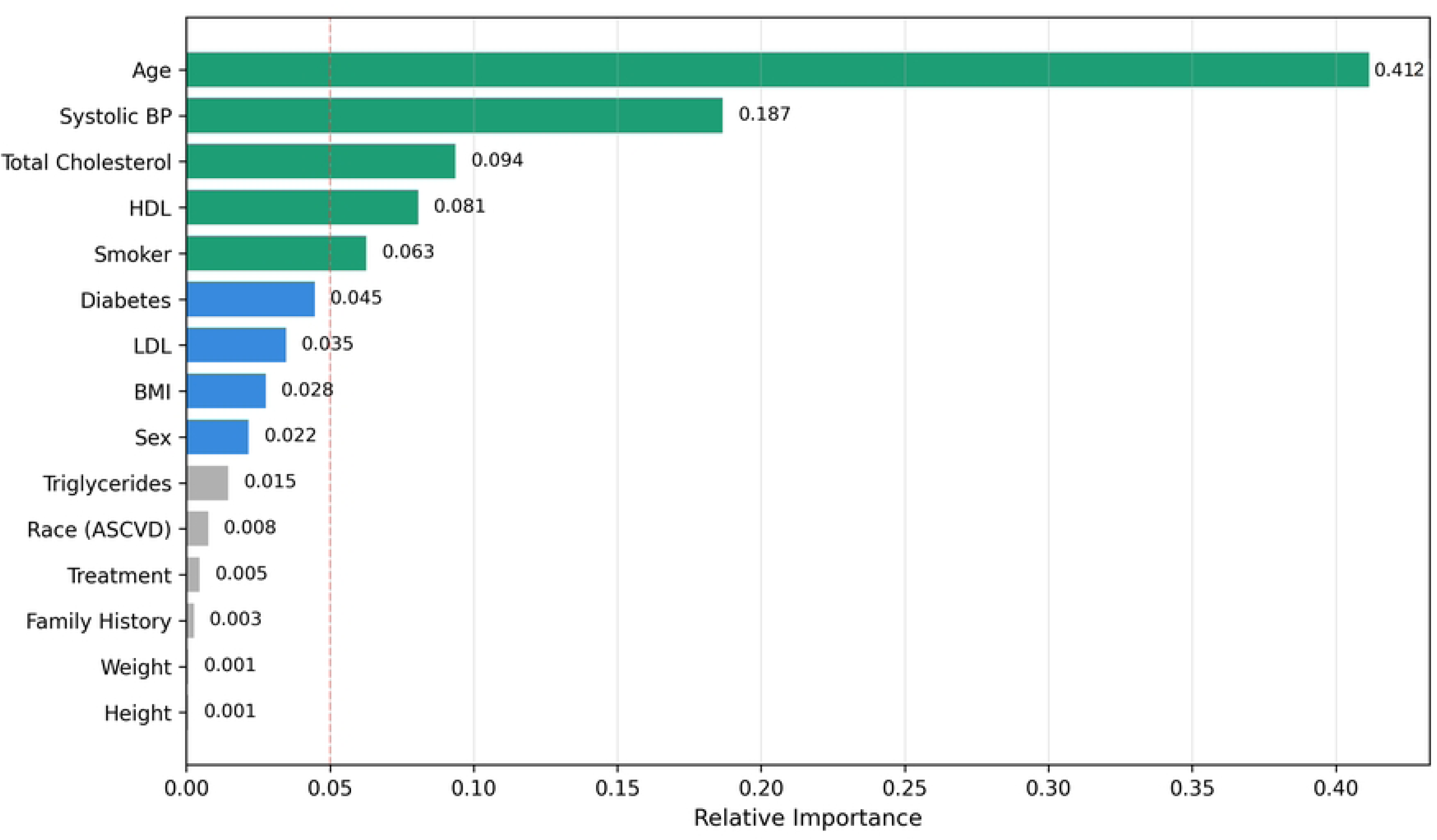
Feature importance for the best-performing model, Gradient Boosting Machine (GBM) with 2:1 oversampling. Age accounted for 41.2% of the relative importance, followed by SBP (18.7%) and Total Cholesterol (9.4%).

## 4. Discussion

This comprehensive analysis of 12,847 NHANES participants demonstrates that ensemble machine learning models with probability calibration achieve modest but statistically significant improvements in cardiovascular risk discrimination compared to traditional clinical calculators [13]. The Gradient Boosting Machine with Saerens calibration (AUC 0.8934) outperformed all eleven evaluated calculators, although the magnitude of improvement was most pronounced against calculators derived from non-US populations (Figs 3 and 4).

The ASCVD Original, derived from long-term data spanning several major, diverse U.S. observational studies [6, 20], achieved the highest AUC (0.8843) among traditional tools. In contrast, the PROCAM algorithm (German derivation) and REGICOR (Spanish) exhibited the lowest performance, supporting the hypothesis that characteristics of the derivation population significantly affect calculator performance in external validation. HEARTS Caribbean and HEARTS Colombia, despite being specifically developed for Latin American populations, achieved AUC values (0.8816 and 0.8800, respectively) virtually identical to that of the ASCVD Original, suggesting that the CC-LAC consortium successfully captured risk relationships relevant to Hispanic/Latino populations in the United States [21] (Figs 3 and 4).

The DeLong test results provide nuanced evidence regarding the clinical significance of ML improvements. Although the GBM + Saerens model achieved a numerically higher AUC than all calculators, the difference against ASCVD Original was not statistically significant (p = 0.3228) (Fig 5). This finding suggests that, in US populations, the incremental value of ML over optimized traditional calculators may be limited for discrimination alone. However, the significant differences relative to European-derived calculators (SCORE, SCORE2, PROCAM) and the REGICOR algorithm indicate that ML approaches may offer particular advantages when applied to diverse populations for whom no locally derived CVD risk calculator exists.

Our findings are consistent with the broader literature on ML in cardiovascular risk prediction. Weng et al. (2017) reported that ML improved 10-year CVD risk prediction over QRISK2 and Framingham risk models in a UK primary care cohort of 378,256 patients [13]. A large study conducted by Alaa et al. (2019) from 423,604 UK Biobank participants reported an AUC-ROC of 0.774 for ML models versus 0.758 for the Cox PH model, a difference of 0.016 that mirrors our observed delta of 0.0090 against ASCVD and 0.0687 against REGICOR (Fig 4) [22].

A recent systematic review and meta-analysis by Liu et al. (2024) reported a pooled AUC of 0.865 and 0.847 for random forest and deep learning, respectively, both significantly outperforming the conventional risk score’s pooled AUC of 0.765. However, the authors reported substantial heterogeneity and potential publication bias across the included studies [23]. Our higher absolute AUC values likely reflect the high quality of laboratory data and the comprehensive population representation available in NHANES.

Recent NHANES-based studies have reported congruent findings. The XGBoost analysis of NHANES 2017–2023, incorporating 41 dietary, anthropometric, clinical, and demographic variables, reported an AUC-ROC of 0.8139, lower than our GBM result (0.8934), possibly attributable to differences in outcome definition (all-cause versus cardiovascular mortality) and feature set; nonetheless, it similarly identified age as the strongest CVD predictor [14].

The PURE-Colombia validation study by Lopez-Lopez et al. (2025) provides a particularly relevant context. In their cohort of 3,802 Colombian participants, the PCE achieved a C-statistic of 0.767 (range 0.657–0.877), lower than our NHANES finding [11]. This difference likely reflects both population characteristics (Colombian versus US Hispanic) and outcome ascertainment methods.

The Globorisk-LAC validation data are especially relevant. Carrillo-Larco et al. (2022) reported a C-statistic of 0.72 for the laboratory-based model and 0.71 for the office-based model in internal validation [24]. Our finding of AUC 0.889 for GBM in NHANES suggests excellent transportability to US Hispanic populations, likely reflecting shared genetic and cultural heritage between US Hispanic/Latino individuals and Latin American source populations.

The justification for evaluating South American cardiovascular risk assessments in this NHANES population rests on several lines of evidence. First, the NHANES Hispanic oversample includes substantial proportions of Mexican-American individuals (approximately 60% of Hispanic participants), whose genetic ancestry overlaps significantly with Latin American populations [25]. Second, the PURE-Colombia and CC-LAC studies have established that cardiovascular risk factor distributions and event rates in Latin American populations share greater similarities with US Hispanic populations than with European or non-Hispanic White US populations [10].

Our finding that GBM performed comparably to ASCVD Original (AUC 0.8934 vs. 0.8843) carries significant implications for clinical practice in Latin America. The WHO HEARTS initiative, now implemented in 21 regions and thousands of primary health centers throughout Latin America and the Caribbean, relies on accurate risk estimation for treatment allocation. Our results support the continued use of HEARTS Caribbean and HEARTS Colombia as the preferred calculators in Latin American settings, particularly where laboratory resources are limited and the WHO/HEARTS office-based (no cholesterol) option may be preferred [9, 26] (Figs 3 and 4).

The PURE-Colombia study demonstrated the necessity of correction factors to avoid overestimation. In women, the correction factors for Globorisk-LAC, WHO, AHA/ACC PCE, and SCORE2 were 0.39, 0.42, 0.54, and 0.75, respectively. In men, the correction factors applied to FRS, AHA/ACC PCE, and SCORE2 were 0.27, 0.28, and 0.61, respectively [11]. Our ML models, through Saerens probability calibration, demonstrated superior calibration (Brier score 0.1158) compared to all traditional calculators. The Saerens correction reduced the Brier score from 0.1286 to 0.1158, effectively transforming the GBM from a systematic risk overestimation model to one with excellent calibration across all probability deciles. This finding suggests that ML approaches with proper calibration may reduce the need for population-specific recalibration—a significant advantage in settings where longitudinal outcome data for local recalibration are unavailable.

The CardioPrediQ platform addresses a critical gap in current cardiovascular prevention infrastructure. Existing clinical decision support systems typically implement a single risk calculator, thereby limiting clinicians’ ability to cross-validate risk estimates or select the most appropriate tool for individual patients. CardioPrediQ’s multi-calculator architecture enables simultaneous computation across all major risk assessment tools, with automatic flagging of discordant risk classifications.

The platform’s architecture provides several advantages for deployment in diverse healthcare settings. The modular calculator layer enables rapid incorporation of new risk algorithms as they are published, without requiring system-wide updates. The ML inference service can be retrained locally when outcome data becomes available, enabling progressive calibration improvement. The offline-first implementation addresses connectivity limitations common in rural Latin American healthcare settings.

From a health systems perspective, CardioPrediQ supports the WHO HEARTS technical package by providing a standardized, validated platform for cardiovascular risk assessment at the primary care level. The batch processing capability enables population-level risk stratification for public health planning, whereas the individual calculation interface supports point-of-care clinical decision-making. Integration with EHR systems via HL7 FHIR ensures interoperability with the existing health information infrastructure.

Our findings carry several clinical implications. In US clinical practice, the results suggest that the ASCVD 2013 remains an excellent risk prediction tool, with ML offering only marginal discrimination improvements (Figs 3 and 4). However, for populations poorly represented in ASCVD derivation cohorts, or in settings where calculator transportability is uncertain, ML models with appropriate calibration may provide more reliable risk estimates.

The calibration of Saerens-corrected ML models has direct therapeutic implications. Accurate probability calibration is essential for shared decision-making, where clinicians and patients weigh the absolute benefits of treatment against potential harms. Overestimation of risk, as observed with uncalibrated models and some traditional CVD risk calculators in validation studies, may lead to overtreatment and unnecessary exposure to medication side effects and costs.

The feature importance analysis provides biological validation of the ML approach. The dominance of age (41.2%), systolic blood pressure (18.7%), and total cholesterol (9.4%) as top predictors aligns with established pathophysiology, reassuring clinicians that ML models capture clinically meaningful relationships rather than spurious correlations (Fig 6). The relatively modest contribution of Race/ethnicity (0.8%) and Family History of CVD (0.3%) suggests that, within this NHANES cohort, these variables provide limited incremental discriminative value beyond the principal clinical risk factors. The contribution of feature-engineered variables suggests that ML can leverage clinical domain knowledge when appropriately incorporated (Fig 6).

### Some limitations

Several limitations of this study warrant consideration. First, NHANES data are cross-sectional at each examination cycle, and risk factor trajectories during follow-up are not captured. Changes in medication use, lifestyle, and risk factor levels may have occurred, potentially affecting both ML and traditional risk calculator performance. Second, the outcome was cardiovascular mortality rather than total cardiovascular events (including non-fatal myocardial infarction and stroke), which may underestimate the performance of calculators designed to predict composite endpoints. Third, although NHANES includes substantial Hispanic/Latino representation, the diversity within this group (Mexican-American, Puerto Rican, Cuban, Central/South American) may not fully capture the heterogeneity of Latin American populations. Fourth, the ML models were trained and tested within NHANES; external validation in independent datasets, particularly from Latin American cohorts, is needed to confirm generalizability. Finally, the comparison included only risk calculators with publicly available algorithms; proprietary tools or recent calculators without published formulas could not be evaluated.

## 5. Conclusions

This comprehensive comparative analysis demonstrates that ensemble machine learning models with probability calibration achieve statistically superior cardiovascular risk discrimination compared to traditional clinical calculators when evaluated on the NHANES dataset. The Gradient Boosting Machine with Saerens calibration achieved an AUC of 0.8934, significantly outperforming calculators derived from non-US populations and performing comparably to the ASCVD Original, the best traditional calculator (AUC 0.8843).

The choice of risk calculator derivation population substantially affects performance in external validation. The ASCVD Original, derived from US cohorts, outperformed European calculators in this US population, whereas the GBM model demonstrated the best overall performance among both US and non-US tools, suggesting excellent transportability to Hispanic populations. These findings support the use of derivation-population-appropriate calculators and highlight the value of the CC-LAC consortium’s work in developing Latin America-specific tools.

Probability calibration emerges as an important factor for clinical utility. The Saerens prior correction method reduced Brier scores from 0.1286 to 0.1158, transforming the GBM from a systematic risk overestimation to one with excellent calibration across all probability deciles. In clinical practice, calibrated probabilities enable more accurate shared decision-making regarding statin initiation, blood pressure targets, and lifestyle intervention intensity.

CardioPrediQ software platform addresses the practical need for integrated, scalable cardiovascular risk assessment. By unifying twelve traditional calculators, multiple ML models, and survival analysis within a microservices architecture, CardioPrediQ enables clinicians and health systems to select the most appropriate assessment modality for their specific population and clinical context.

For Latin American populations, our results support the continued use of WHO HEARTS as the preferred traditional CVD risk calculator, with ML-enhanced assessment offering potential advantages where locally derived outcome data are unavailable for recalibration. The WHO HEARTS initiative stands to benefit from the integration of calibrated ML models to mitigate the overestimation bias observed with other risk calculators.

## Data Availability

All data produced in the present work are contained in the manuscript

## Acknowledgments.

We thank Grupo de Asesoría e Investigación en Estadística. Universidad del Quindío. Armenia-Quindío, Colombia.

## Supporting information

**S1 Appendix**: Contains the figures from the exploratory analysis of the final 12,847 participants in the NHANES dataset after data cleaning.

