## Supplementry material for "Comparative Analysis of Machine Learning Models vs. Traditional Clinical Calculators for Cardiovascular Risk Prediction"

### S1 Appendix

**Participant flow through the nine-stage cleaning pipeline. The nine stages were described in the Materials and Methods section.**

| Stage | Criterion | n Total | Alive (%) | Deceased (%) | Eliminated |
|---|---|---|---|---|---|
| 0 | Raw NHANES data | 230,000 | – | – | 0 |
| 1 | Race/ethnicity filter | 80,932 | 37,017 (45.7) | 13,262 (16.4) | 50,098 |
| 2 | Missing clinical variables | 43,374 | 28,931 (66.7) | 14,443 (33.3) | 37,558 |
| 3 | Follow-up/cause filter | 20,462 | 17,524 (85.6) | 2,938 (14.4) | 22,912 |
| 4 | SEQN deduplication | 19,025 | 16,087 (84.6) | 2,938 (15.4) | 1,437 |
| 5 | Smoker NaN elimination | 17,913 | 15,131 (84.5) | 2,782 (15.5) | 1,112 |
| 6 | Age range (30–80) | 13,573 | 11,448 (84.3) | 2,125 (15.7) | 4,340 |
| 7 | Clinical range filters | 13,501 | 11,394 (84.4) | 2,107 (15.6) | 72 |
| 8 | Physiological filters | 12,850 | 10,851 (84.4) | 1,999 (15.6) | 651 |
| 9 | IQR × 3 outliers | 12,847 | 10,805 (84.1) | 2,042 (15.9) | 3 |

### Age Distribution

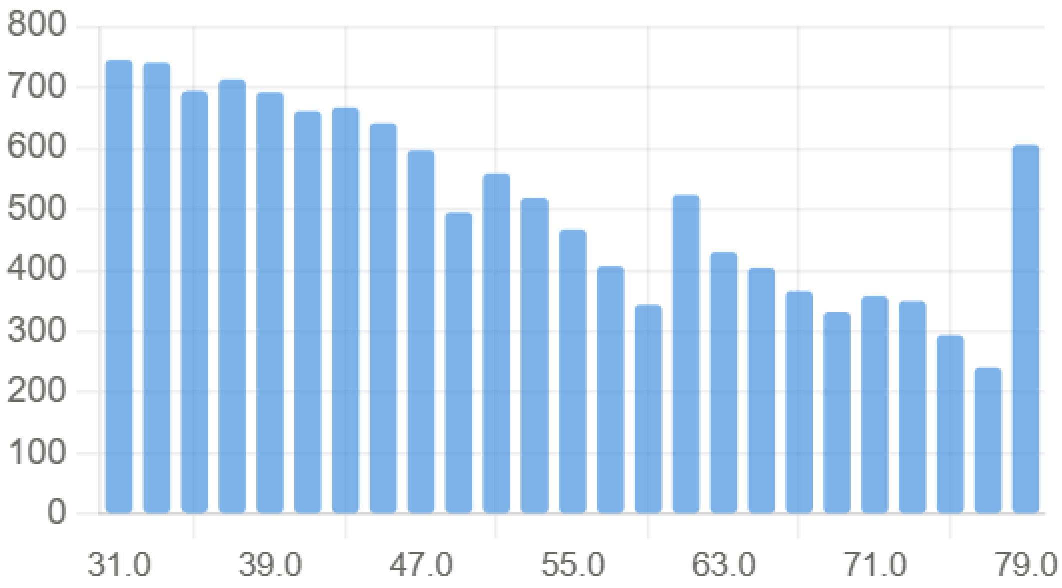

**Genre Distribution.**

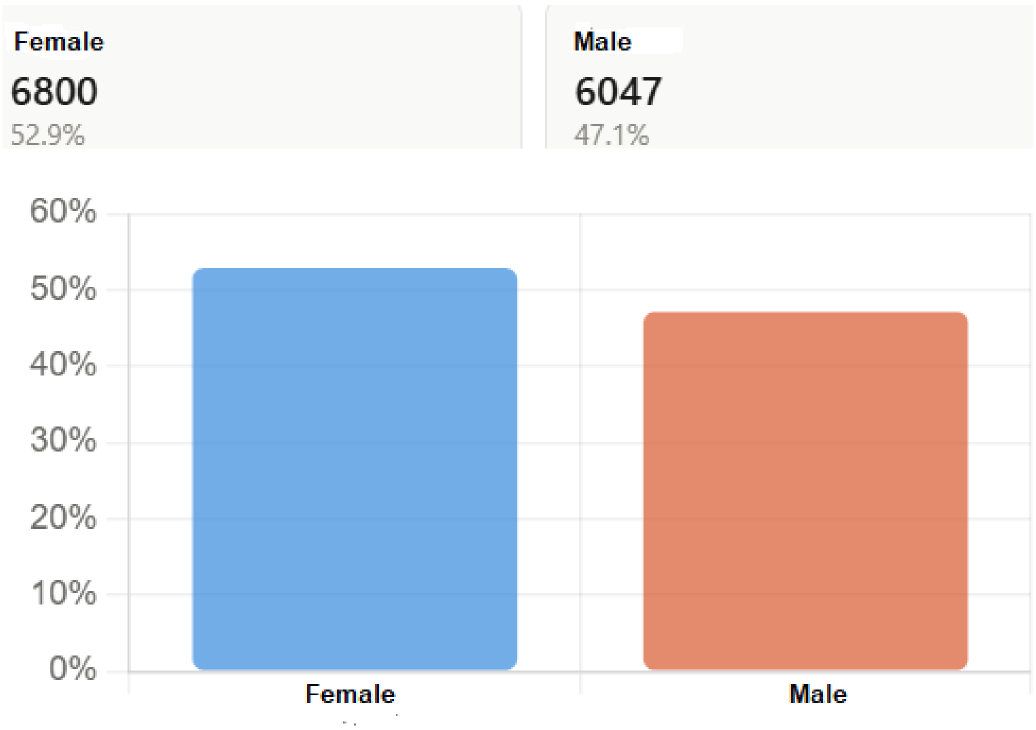


**Race/ethnicity Distribution. White includes Hispanic/Latino and North American white. Black is non-Hispanic Black (AfroAmerican). This variable is necessary for ASCVD risk calculation.**

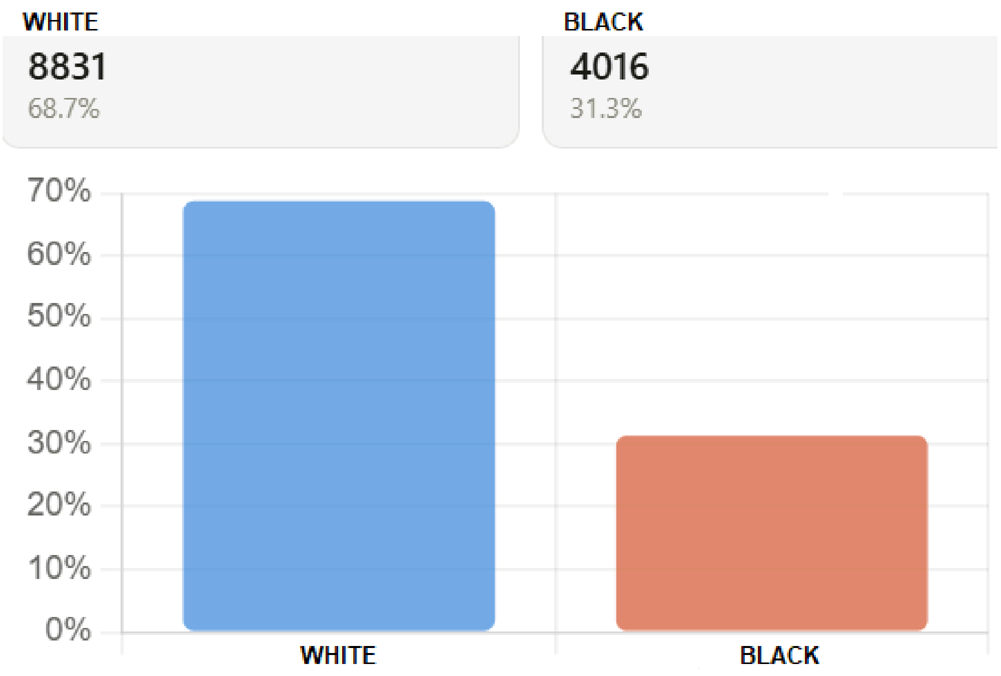

### Hypertension (HTA) Treatment Distribution.

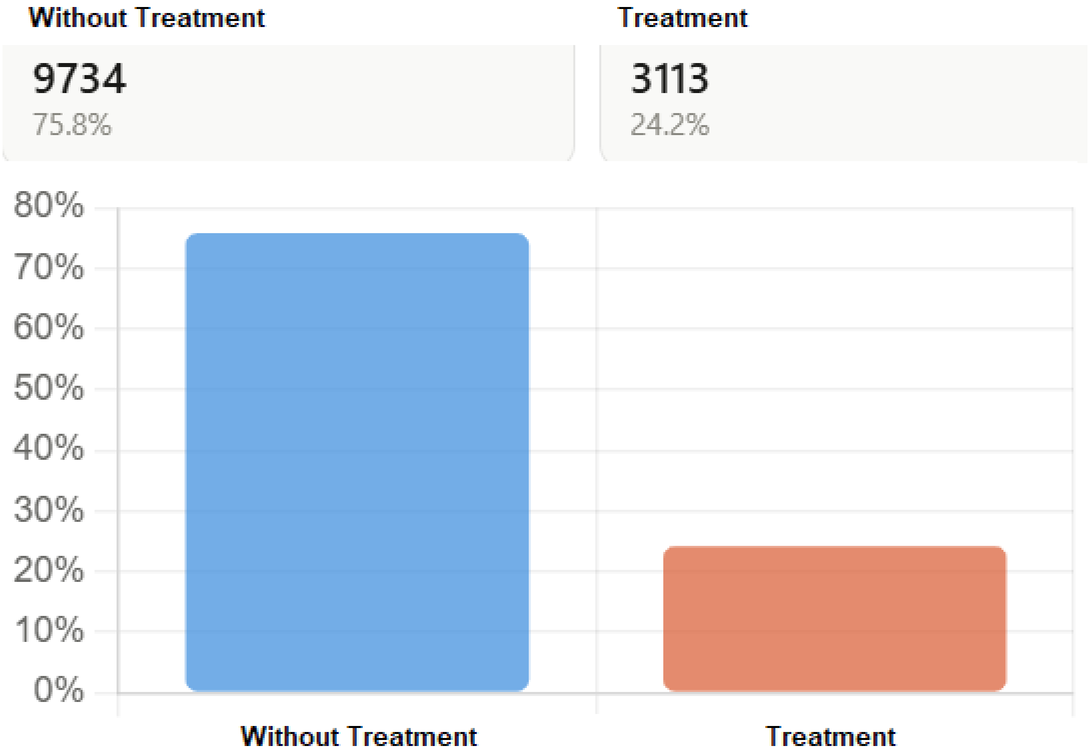


### Smoking Distribution.

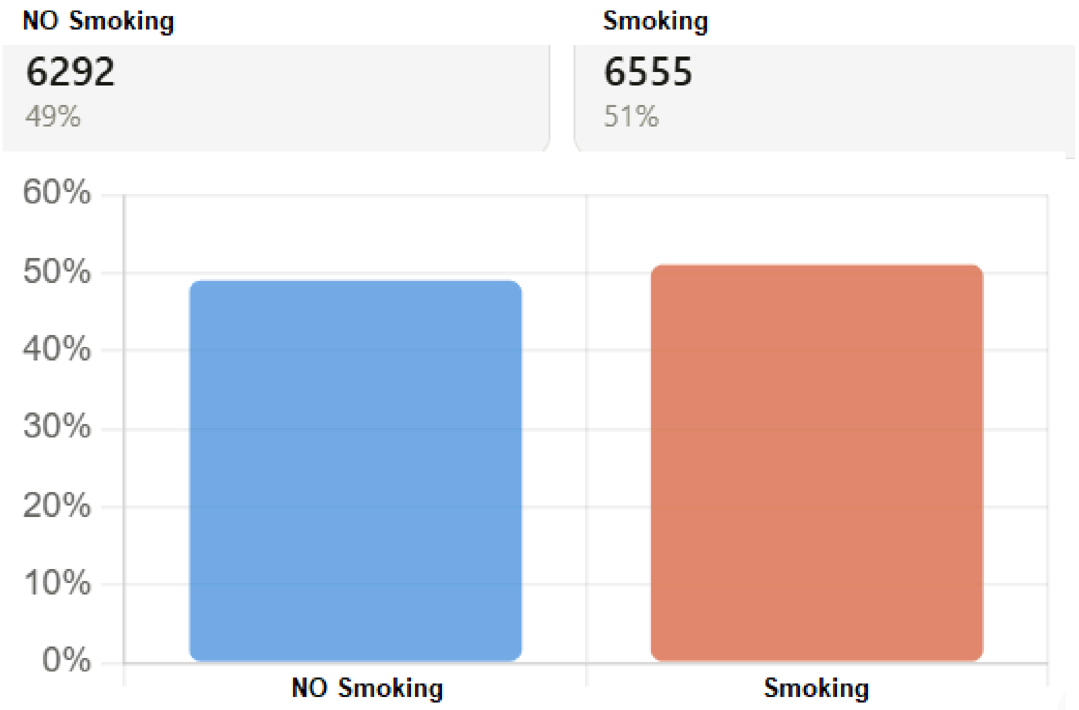

**Diabetes Distribution**

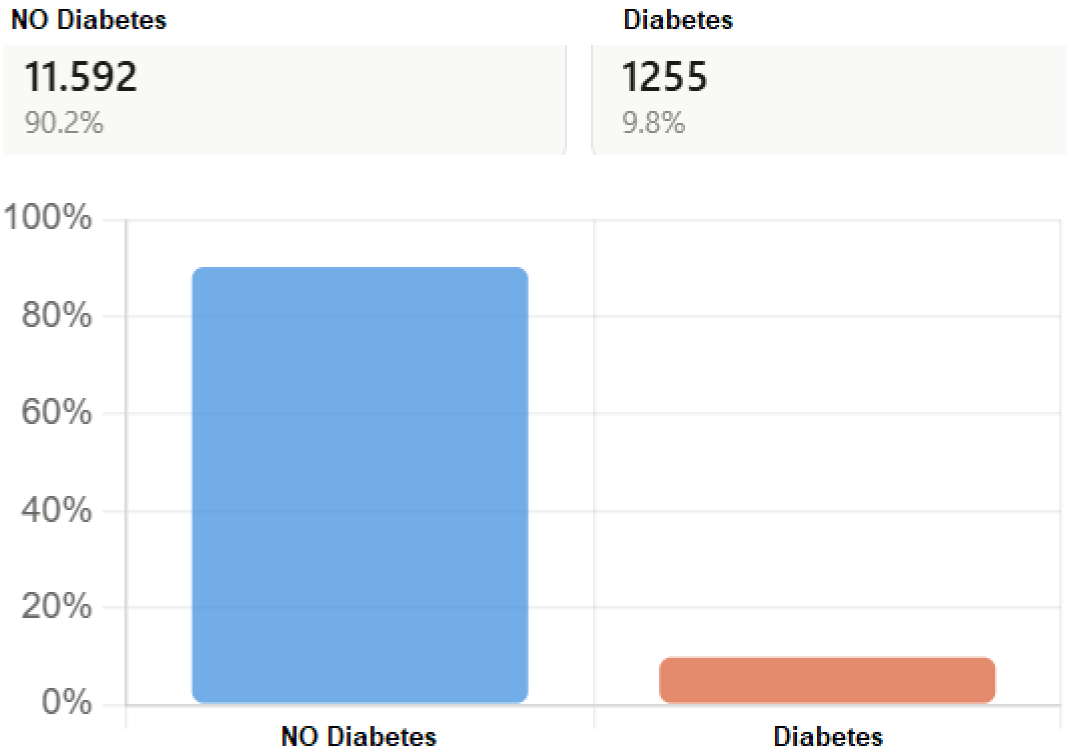


**Distribution of the family history with premature cardiovascular disease (FHPCVD).**

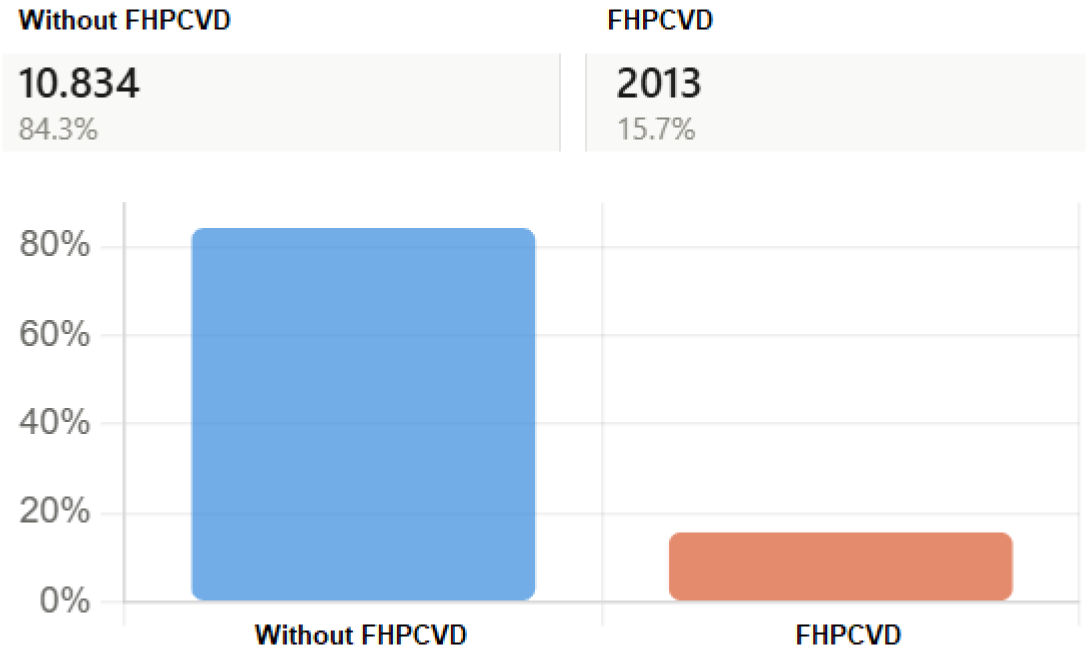

**Survival Status Distribution  (Alive and Deceased)**

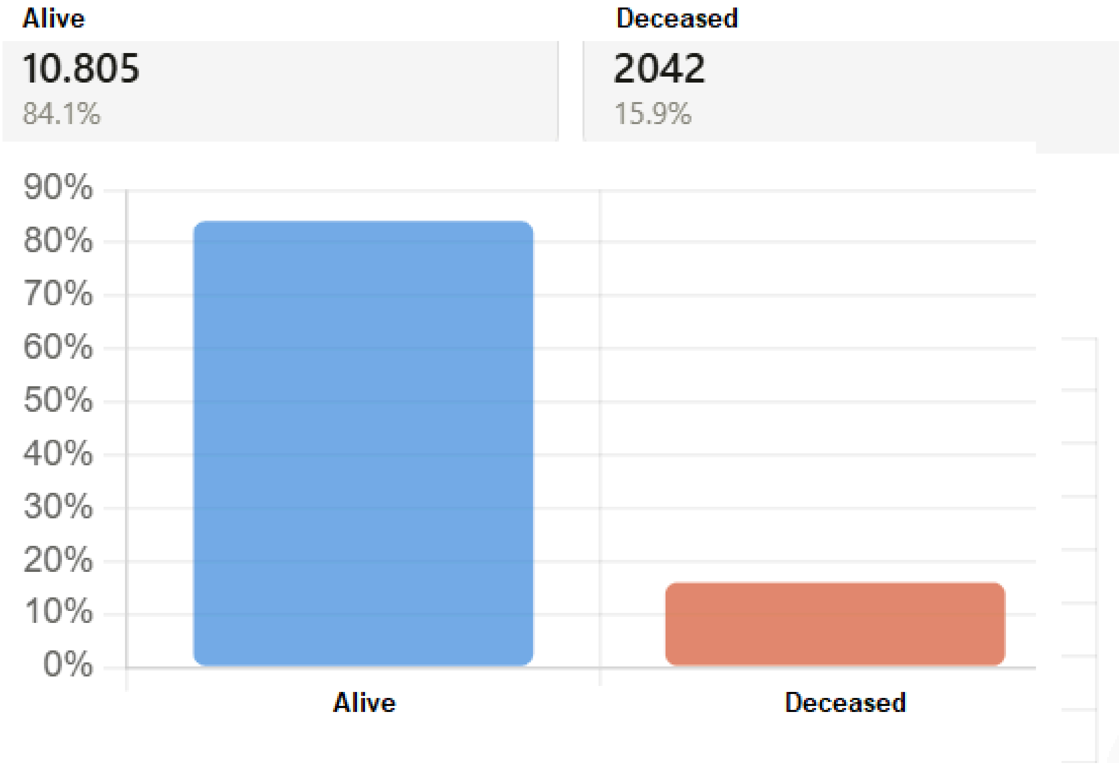


**Variable Description. Age in years; SBP (Systolic Blood Pressure); Weight in Kilograms; Height in centimeters; Total cholesterol, HDL, LDL, and Triglycerides were taken in mg/dL. SD means standard deviation.**

| Variable | Mean | SD | Min | Max |
|---|---|---|---|---|
| AGE | 50.87 | 14.48 | 30 | 80 |
| SBP | 125.13 | 17.85 | 90 | 180 |
| Total Cholesterol | 204.41 | 40.55 | 100 | 390 |
| HDL | 53.63 | 15.94 | 20 | 125 |
| LDL | 123.52 | 36.56 | 20.6 | 289 |
| Triglycerides | 131.75 | 73.13 | 30 | 400 |
| Weight | 82.65 | 19.39 | 32.05 | 168.8 |
| Height | 170.76 | 9.42 | 135.4 | 206.5 |

**Age Distribution by Survival Status. P25 and P75 refer to percentiles 25 and 75, respectively.**

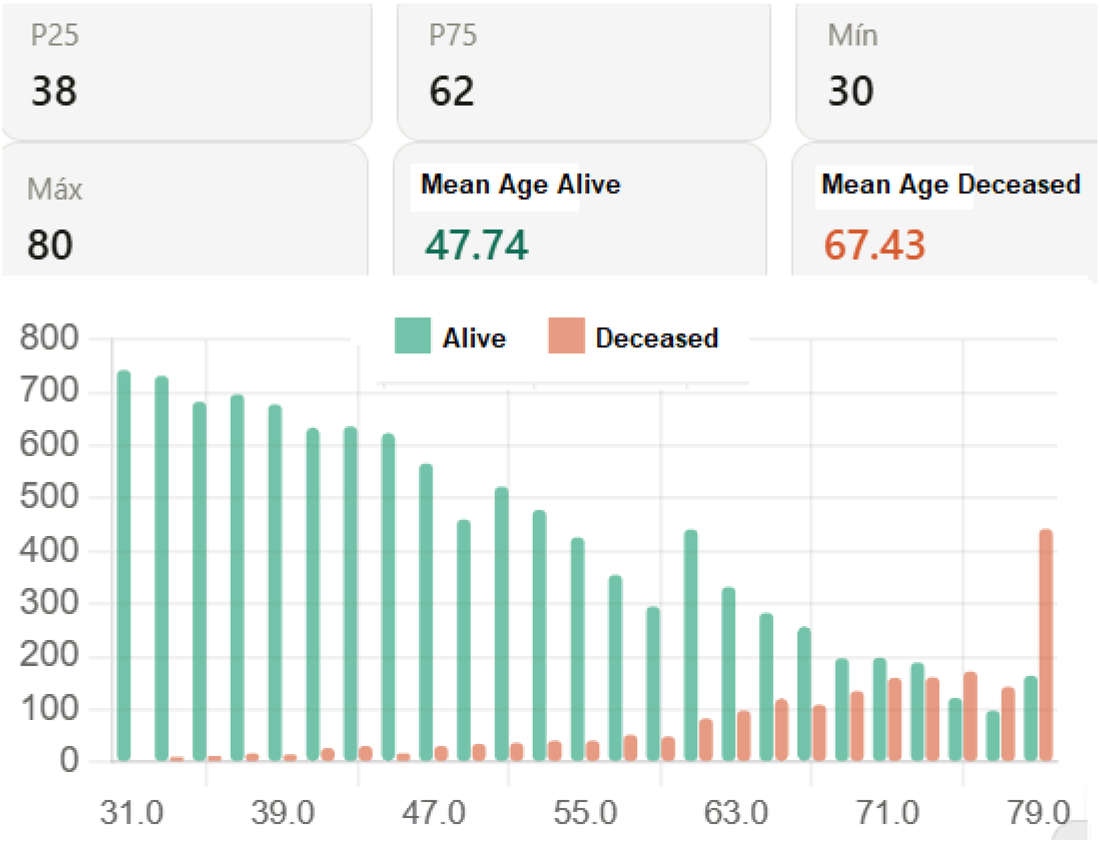
